# Anatomy-Aware 3D GAN with Local Mask Refinement for Brain MRI Inpainting

**DOI:** 10.64898/2026.11.12.26367904

**Authors:** Abdul Fatah Rahimi, Hari Mohan Rai, Sayed Yaser Ahmad Mushtaq

## Abstract

Brain tumor MRI inpainting aims to synthesize plausible healthy-appearing tissue in regions removed from pathological scans. We present an anatomy-aware three-dimensional generative adversarial network for Task 4 of the BraTS 2026 challenge. The method combines mask-aware gated convolutions, mirrored contralateral context, attention-modulated skip fusion, boundary-aware refinement, and a local masked-region refiner, each adapted from established components in the image synthesis and inpainting literature. The model is trained with a two-stage objective that first emphasizes voxel-wise and structural reconstruction and then applies low-weight adversarial fine-tuning. On a deterministic local held-out split, the proposed method substantially improves over a simple 3D AutoEncoder baseline but does not surpass the challenge’s Pix2Pix3D baseline. On the official BraTS 2026 Task 4 evaluation server, the submitted model obtained a mean SSIM of 0.658 and a mean PSNR of 14.81 dB over 219 evaluated cases, lower than the local held-out figures. We report these results transparently, analyze likely contributing factors, and identify the controlled ablations (removal of the mirrored-context channel and of the two-stage schedule) required to isolate the contribution of each architectural component.

## 1 Introduction

The Brain Tumor Segmentation (BraTS) challenge has become a central bench-mark for neuro-oncology image analysis, initially focusing on glioma segmentation and later expanding toward multiple tumor entities, clinical settings, and image synthesis tasks [1-3]. Task 4 of the BraTS 2026 challenge addresses local synthesis or inpainting of brain tumor MRI scans, where the goal is to reconstruct healthy-appearing tissue in regions removed from a pathological T1-normalized image. This task is motivated by the need to make tumor-bearing scans more compatible with algorithms developed for healthy or non-lesional brain images, including parcellation, tissue segmentation, image registration, and downstream morphometric analysis [4].

Medical image inpainting differs from natural image completion in several important ways. First, the result must be anatomically plausible in three dimensions rather than only visually coherent in a two-dimensional projection. Second, output intensities must remain consistent with the original scan so that the synthesized region does not introduce artificial discontinuities. Third, the prediction must not alter the known non-masked tissue, because unnecessary changes outside the target region may invalidate the output for downstream analysis. Finally, the evaluation region is explicitly defined by a binary mask, making the challenge objective highly localized.

The challenge baseline includes a 3D AutoEncoder and a 3D Pix2Pix-style model. Pix2Pix introduced conditional adversarial learning for paired image-to-image translation and remains a strong reference for synthesis problems [5]. However, direct adversarial optimization can sometimes reduce voxel-wise similarity metrics when the discriminator encourages visually plausible but numerically shifted outputs. For the inpainting challenge, where metrics include structural similarity index measure (SSIM), peak signal-to-noise ratio (PSNR), mean squared error (MSE), and root mean squared error (RMSE), reconstruction accuracy remains central [6].

We investigate AnatomyAwareGAN3D, a three-dimensional inpainting model that combines the binary mask, approximate left-right brain symmetry, boundary supervision, and local masked-region refinement into a single architecture. Rather than proposing new individual components, our contribution lies in adapting and combining several established techniques from the general image-synthesis literature — gated convolutions [7], attention-gated skip connections [8], residual learning [9], and spectral normalization [10] — for the specific constraints of masked 3D MRI inpainting, where only a small, explicitly defined sub-volume is evaluated. We evaluate this combination on a local held-out split against the challenge’s own AutoEncoder and Pix2Pix3D baselines and report the result candidly, including that the method does not yet close the gap to Pix2Pix3D. We also report the score returned by the official challenge evaluation server for our submitted container.

Our contributions are as follows:

– We adapt gated convolutions, mirrored contralateral context, attention-modulated skip fusion, boundary refinement, and local masked-region refinement — each grounded in prior work —into a single 3D generator for mask-constrained MRI inpainting.
– We describe a two-stage training strategy that combines masked L1, SSIM, multi-scale masked L1, boundary-ring loss, and low-weight adversarial fine-tuning, and we report checkpoint selection by two different validation criteria.
– We provide a compact Docker inference pipeline that preserves the original non-masked tissue and writes challenge-compliant NIfTI outputs.
– We evaluate the model on a deterministic local held-out split, compare against AutoEncoder and Pix2Pix3D baselines, report the official server-side score of the submitted model, and explicitly identify the ablations needed to attribute performance to individual architectural choices, which remain as future work.

## 2 Related Work

### 2.1 Brain Tumor MRI Benchmarks

BraTS has played a major role in standardizing brain tumor MRI analysis by providing curated multimodal datasets, benchmark tasks, and common evaluation protocols [1, 2]. The 2026 challenge extends this tradition through multiple tasks that address segmentation, generalizability, pathology-related prediction, and local synthesis [3]. The inpainting task is especially relevant because tumor-bearing anatomy often breaks assumptions made by tools developed for healthy subjects.

### 2.2 Medical Image Inpainting

The BraTS local synthesis task was introduced to encourage methods that can replace tumor-affected or removed tissue with plausible healthy tissue [4]. Unlike segmentation, where the output is a label map, inpainting requires voxel-level synthesis of a continuous intensity image. This makes the task sensitive to intensity normalization, boundary blending, and contextual consistency. Strong inpainting models must infer missing anatomy while preserving the identity of the scan outside the mask.

### 2.3 Generative Models for Image-to-Image Translation

Generative adversarial networks (GANs) introduced a framework for training generators using learned discriminators [11]. Conditional GANs, including Pix2Pix, extended this idea to paired image-to-image translation [5]. Least-squares GANs improved stability by replacing binary cross-entropy with a regression-style adversarial objective [12]. Our model uses a conditional adversarial loss only during fine-tuning and keeps the adversarial weight small, because the target challenge metrics reward similarity to hidden ground truth rather than unrestricted perceptual realism.

### 2.4 Architectural Components Adapted in This Work

The generator and discriminator components used in this paper are individually well established, and we position each explicitly rather than presenting them as novel. Gated convolutions were introduced for free-form image inpainting to let the network learn a dynamic, per-location feature gate conditioned on the mask, which also mitigates the color-discrepancy and blurry-boundary artifacts common to plain-convolution inpainting networks [7]; we adopt this mechanism directly for the 3D setting. Attention-gated skip connections were proposed in

Attention U-Net to suppress irrelevant encoder activations before they are fused into the decoder, originally for pancreas segmentation [8]; we reuse the same gating principle, conditioning the attention map additionally on the inpainting mask so that skip features near the voided region are down-weighted unless the decoder attends to them. Residual connections follow the formulation of He et al. [9] and are used throughout the bottleneck to stabilize gradient flow in a deep 3D network. The discriminator uses spectral normalization [10] to bound its Lipschitz constant and stabilize adversarial fine-tuning. U-Net-style encoder-decoder architectures with skip connections [13] and their 3D extension [14] form the backbone on which these components are assembled. The novelty of our approach is therefore the specific combination and mask-conditioning of these established components for localized 3D MRI inpainting, not the individual mechanisms themselves.

Regarding the mirrored-context channel, approximate left-right structural symmetry of the human brain is a documented, though imperfect, anatomical property: healthy brain hemispheres show strong but not exact correspondence, with well-characterized regions of structural and functional asymmetry [15]. We use this property only as a weak prior — an additional input channel the network may draw on — rather than as an assumption enforced on the output, and we discuss its limitations under pathology in Sect. 5; we treat this as an open question rather than a demonstrated result (see Sect. 4.3 and Sect. 5).

## 3 Methods

### 3.1 Task Definition

Each inference case consists of a voided T1-normalized MRI volume and a binary mask. The model receives the pair

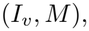

where *I*_*v*_ is the voided image and *M* indicates the region to be synthesized. The objective is to produce a full-resolution image *I*_*o*_ such that

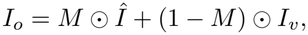

where *Î* is the network prediction and⊙ denotes element-wise multiplication. This formulation ensures that the known non-masked region is preserved exactly. It also reduces the risk of invalid predictions by constraining the model to affect only the region requested by the challenge.

Figure 1 illustrates the task inputs and intended output schematically.

**Fig. 1.**
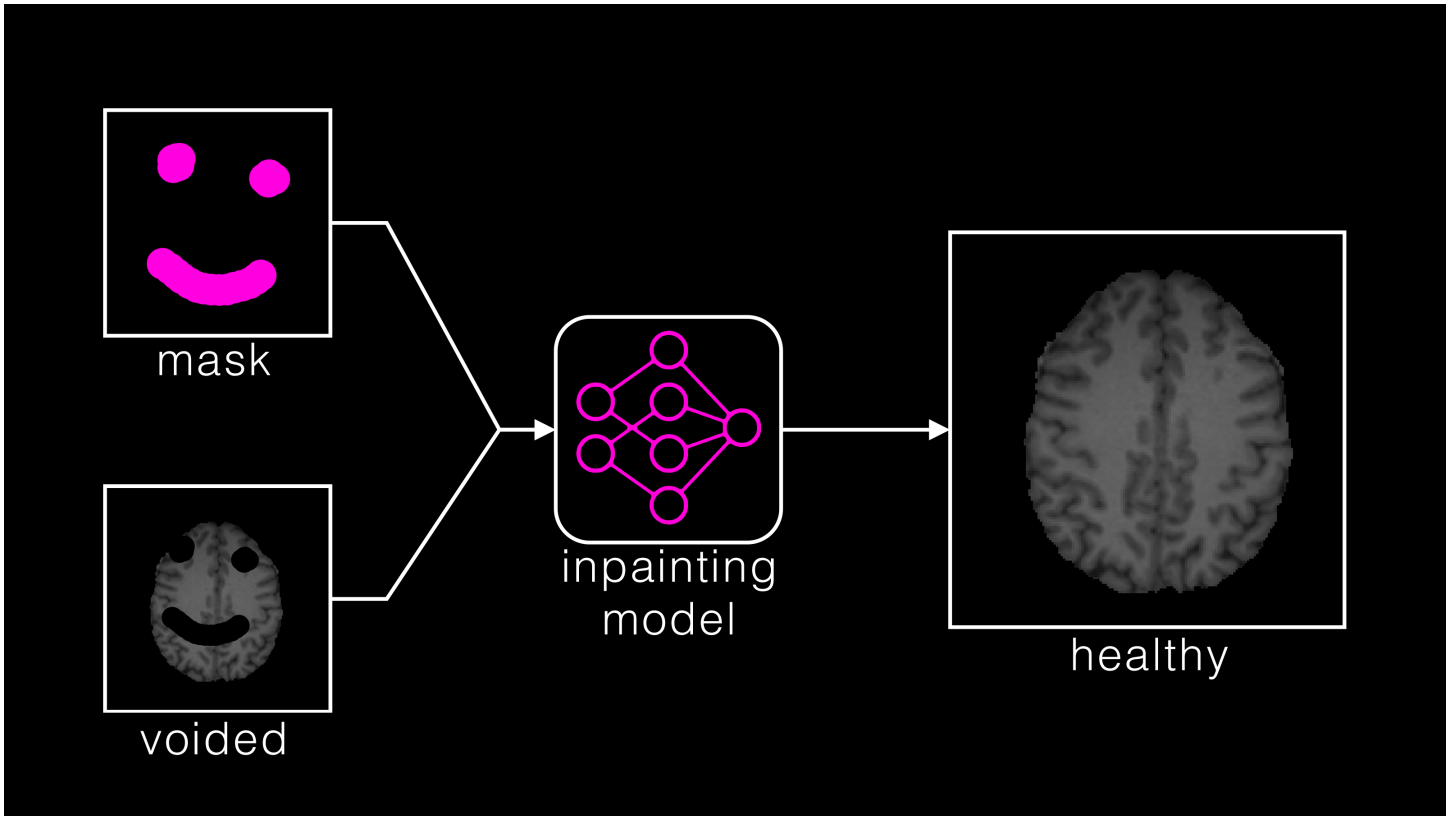
MRI inpainting task schematic, reproduced from Kofler et al. [4] (their Fig. 1). A binary mask (magenta) identifies missing regions in the voided image; an inpainting model uses both inputs to synthesize healthy-appearing tissue. The “healthy” output label denotes the intended appearance, not verified healthy anatomy. The stylized mask illustrates the task and is not a tumor annotation or a prediction example from our evaluation.

### 3.2 Preprocessing

We follow the preprocessing utilities provided in the challenge baseline repository. Each volume is clipped to remove negative intensity noise and normalized to [0, 1]. During training, the target T1 image and healthy-mask label are loaded, and a voided image is generated by masking out the target region. During inference, the provided voided image and mask are loaded directly. Our local experiments use 1251 training cases and 219 held-out validation cases from the BraTS 2026 inpainting training release, split deterministically with a fixed random seed (Sect. 4.1).

The mask bounding box is used to define a centered three-dimensional crop of fixed size

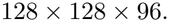

This size was chosen empirically as the smallest fixed box that contained the large majority of mask bounding boxes observed during development while remaining within the memory budget of a single training GPU; masks whose bounding box exceeds this size are still fully covered because the network operates on the full crop rather than only the mask interior, but the surrounding context available to the model is correspondingly reduced. If a crop extends beyond the image boundary or is smaller than the target shape, padding is applied. Cropped images are mapped to [−1, 1] prior to being passed to the network; this range matches the output activation of the generator’s final tanh layer (Sect. 3.5) and is the standard convention for GAN-based image synthesis, avoiding the saturation and gradient-scaling issues that arise when a bounded output activation must match an unbounded or [0, 1]-scaled target. At inference time, the predicted crop is restored to full image space and rescaled to the original intensity range. The affine matrix from the input NIfTI file is preserved, which is required for challenge compliance and for any downstream use of the produced image.

### 3.3 Generator Architecture

The generator is a three-dimensional encoder-decoder network. Its input contains three channels:

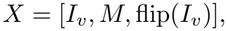

where flip(*I*_*v*_) is the left-right mirrored version of the voided image, included as a weak prior toward approximate hemispheric symmetry (Sect. 2.4). The model architecture is summarized in Fig. 2.

**Fig. 2.**
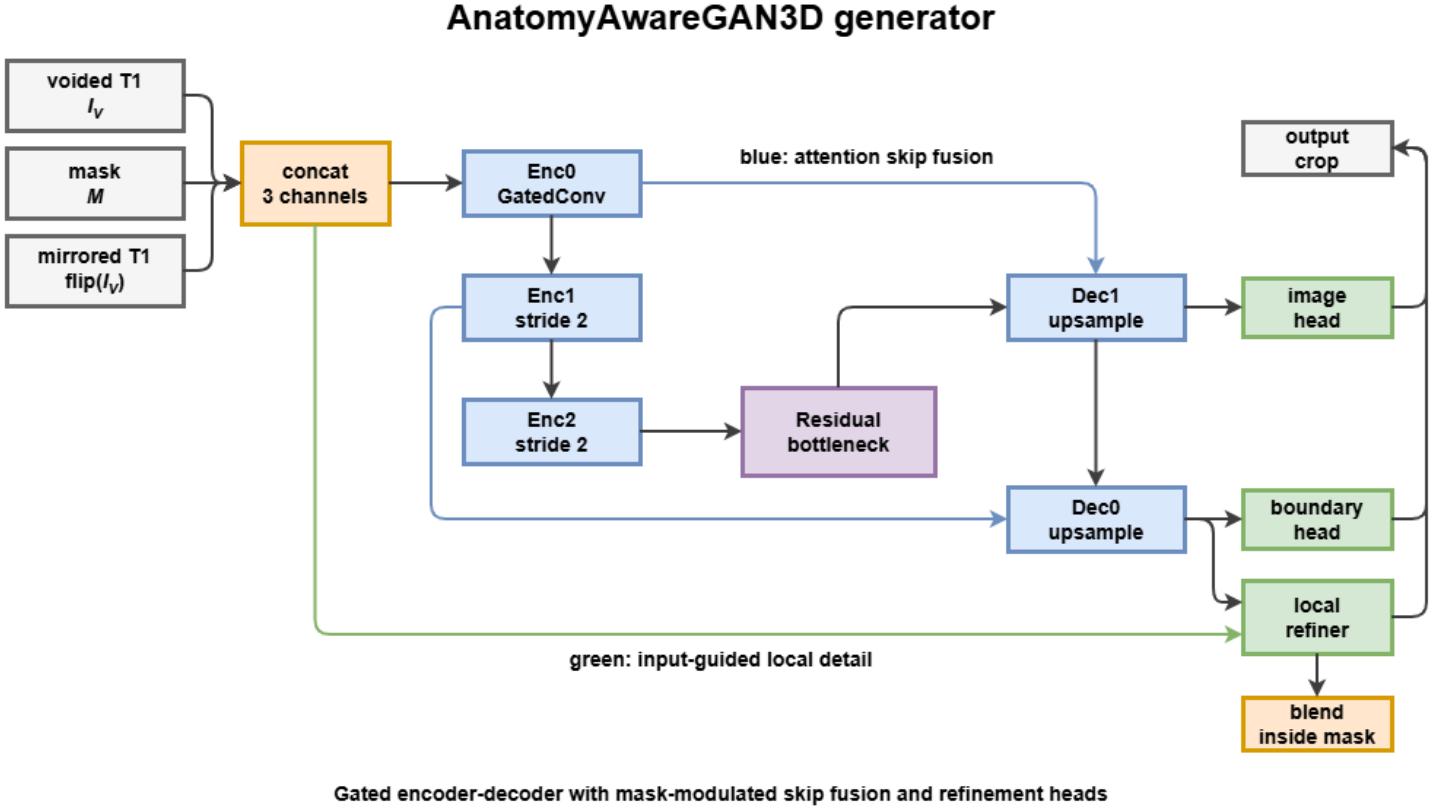
Overview of the AnatomyAwareGAN3D generator. The network receives the voided T1 image, binary mask, and mirrored voided image. Gated convolutional encoding and residual bottleneck features are decoded through attention-modulated skip fusion. Boundary and local refinement heads focus synthesis on the mask and its transition region.

The encoder uses gated 3D convolution blocks following the formulation of Yu et al. [7], adapted to 3D. A gated convolution computes both a feature tensor and a learned gate:

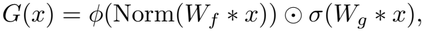

where *W*_*f*_ and *W*_*g*_ are convolutional kernels, *ϕ* is the GELU activation, and *σ* is the sigmoid function. This mechanism allows the model to modulate information flow based on local context and the mask, rather than treating masked and valid voxels identically as a plain convolution would. The bottleneck consists of residual gated convolution blocks in the style of He et al. [9], which stabilizes training and supports deeper context aggregation. The decoder upsamples latent features using trilinear interpolation followed by gated convolution.

### 3.4 Attention-Modulated Skip Fusion

Skip connections are important for preserving spatial detail, but direct skip copying can also reintroduce voided-region content into the decoder, a known failure mode in mask-conditioned generators [7]. We address this using an attention gate adapted from Attention U-Net [8], additionally conditioned on the mask. For a skip tensor *S* and decoder tensor *D*, the attention map is computed from their concatenation with a resized mask:

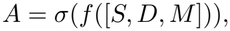

and the fused skip features are obtained by multiplying *S* with *A*. Because *A* is conditioned on *M*, the network can learn to down-weight skip features inside or near the voided region while passing valid-tissue features through largely unmodified, rather than treating all encoder features uniformly as in a standard skip connection.

### 3.5 Boundary and Local Refinement

Boundary artifacts at the mask edge are a well-documented failure mode in inpainting, arising because the generated region must blend seamlessly into known tissue [7]. We address this with a boundary refinement head that predicts a small residual correction restricted to a band around the mask boundary; the residual is scaled by a small learned factor before being added to the main prediction, preventing unstable intensity shifts at the transition.

The final model also includes a local masked-region refiner, shown in Fig. 3. Because the challenge metrics are computed only inside *M* (Sect. 4.1), errors outside the mask do not affect scoring, motivating a branch that concentrates additional network capacity on the interior of the mask specifically. This branch receives decoder features concatenated with the original input channels and predicts a local residual and a learned blending map. The residual is applied only inside the binary mask:

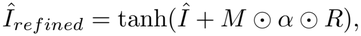

**Fig. 3.**
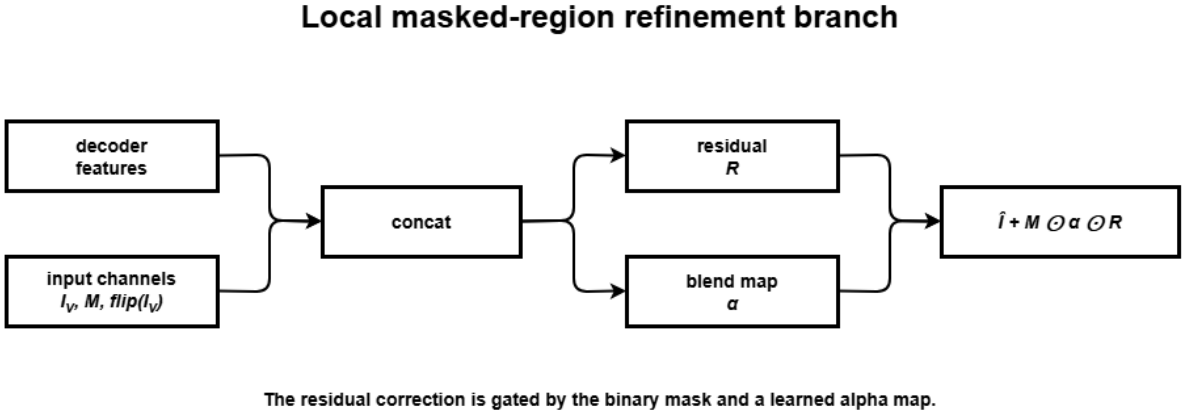
Local masked-region refiner. Decoder features and original conditional inputs are fused to predict a residual correction and learned blend map. The correction is gated by the binary mask.

where *R* is the local residual and *α* is the learned blend map.

### 3.6 Discriminator

The discriminator is a 3D PatchGAN-style network [5] with spectral normalization applied to every convolutional layer [10], which bounds the discriminator’s Lipschitz constant and reduces the risk of destabilizing gradients during adver-sarial fine-tuning. It receives a concatenation of the synthesized or real image with the conditional input. The discriminator is used only during the second training stage with a small adversarial weight, since strong adversarial loss may reduce reconstruction metrics.

### 3.7 Loss Functions

The generator is optimized with a weighted combination of losses. The primary loss is masked L1 reconstruction:

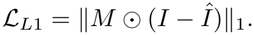

We also use SSIM loss [6]:

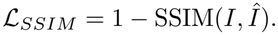

SSIM is computed over the full crop rather than restricted to *M* : SSIM is a windowed statistic that compares local means, variances, and covariances within a sliding kernel, and applying it only to the irregular interior of a binary mask would require ad hoc handling at the window boundary and would discard the local-contrast information from immediately adjacent valid tissue that SSIM depends on. Because the crop is centered on the mask (Sect. 3.2), the masked region dominates the crop and still receives the great majority of gradient signal from this term. A multi-scale masked L1 loss is computed after average pooling by factors of 2 and 4, restricted to *M* at each scale, encouraging both fine local reconstruction and coarse anatomical consistency. A boundary-ring loss is computed on a band formed by mask dilation and erosion:

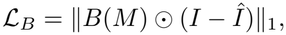

where *B*(*M* ) is the boundary band. During adversarial fine-tuning, we add a least-squares GAN loss [12] with a small weight.

The final generator loss is:

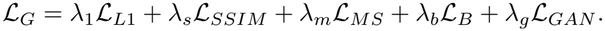

### 3.8 Training Protocol

Training uses an 80/20 deterministic split with seed 2023. The model is trained in two stages. Stage 1 is reconstruction-focused and disables adversarial learning:

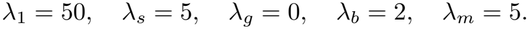

Stage 2 resumes from the best Stage 1 checkpoint and enables low-weight adver-sarial fine-tuning:

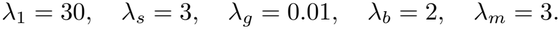

Both stages use a boundary kernel size of 9. We track two independent checkpoint-selection criteria throughout training - lowest validation reconstruction loss and highest validation SSIM - and report both resulting models in Table 1 (rows labeled by selection criterion), since they represent different, non-identical trade-offs rather than a single “best” model (Sect. 4.3). We emphasize that both criteria are computed on the same held-out training-data split used throughout this paper, not on the challenge’s hidden validation or test sets; the selected checkpoint therefore reflects the model that generalizes best within our local split and is not itself evidence of generalization to the official hidden data, which can only be established by the server-side evaluation (Sect. 4.4). The two-stage optimization flow is summarized in Fig. 4.

**Table 1.**
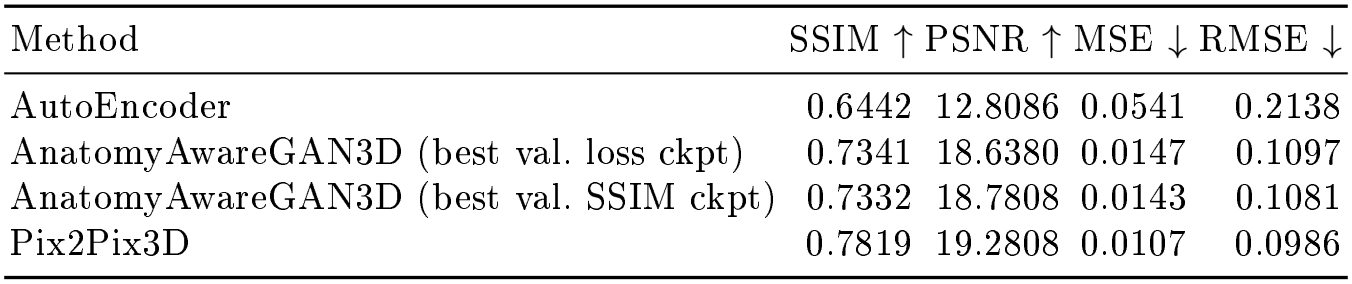
Local held-out results on the deterministic validation split. Metrics are computed inside the inpainting mask. The two AnatomyAwareGAN3D rows are the same architecture and training run, evaluated at two different checkpoints selected by the two criteria described in Sect. 3.8; they are not independent models.

| Method | SSIM ↑ | PSNR ↑ | MSE ↓ | RMSE ↓ |
|---|---|---|---|---|
| AutoEncoder | 0.6442 | 12.8086 | 0.0541 | 0.2138 |
| AnatomyAwareGAN3D (best val. loss ckpt) | 0.7341 | 18.6380 | 0.0147 | 0.1097 |
| AnatomyAwareGAN3D (best val. SSIM ckpt) | 0.7332 | 18.7808 | 0.0143 | 0.1081 |
| Pix2Pix3D | 0.7819 | 19.2808 | 0.0107 | 0.0986 |

**Fig. 4.**
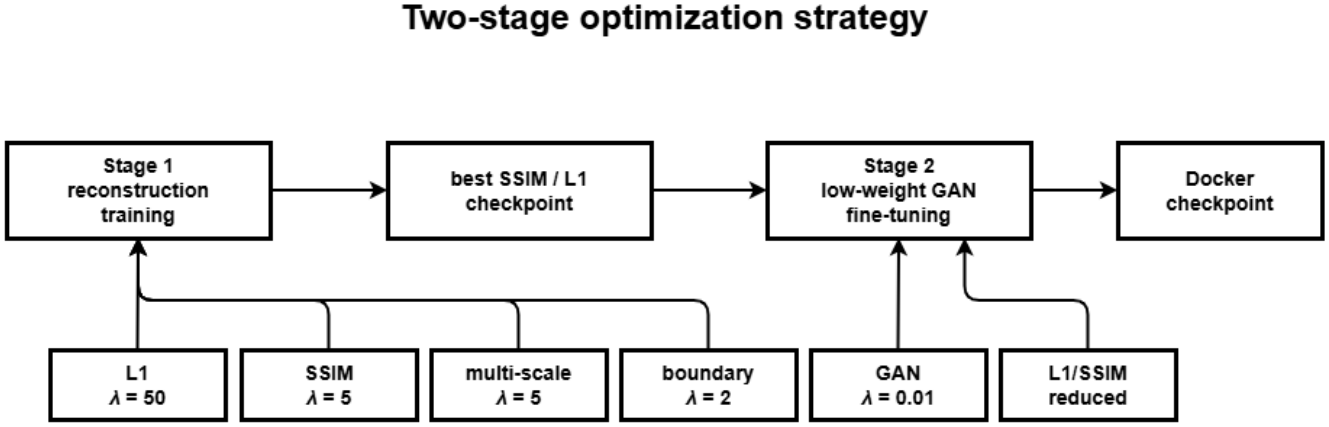
Two-stage training strategy. Stage 1 emphasizes reconstruction losses, while Stage 2 resumes from the best checkpoint and introduces a small adversarial term.

### 3.9 Deployment

For challenge submission, the trained model is packaged in a CUDA-enabled Docker image that applies the preprocessing of Sect. 3.2 to each input case, runs inference, restores the prediction to full image space, and writes one NIfTI file per case with non-masked voxels copied directly from the input voided image, matching the formulation in Sect. 3.1. Figure 5 summarizes the inference pipeline.

**Fig. 5.**
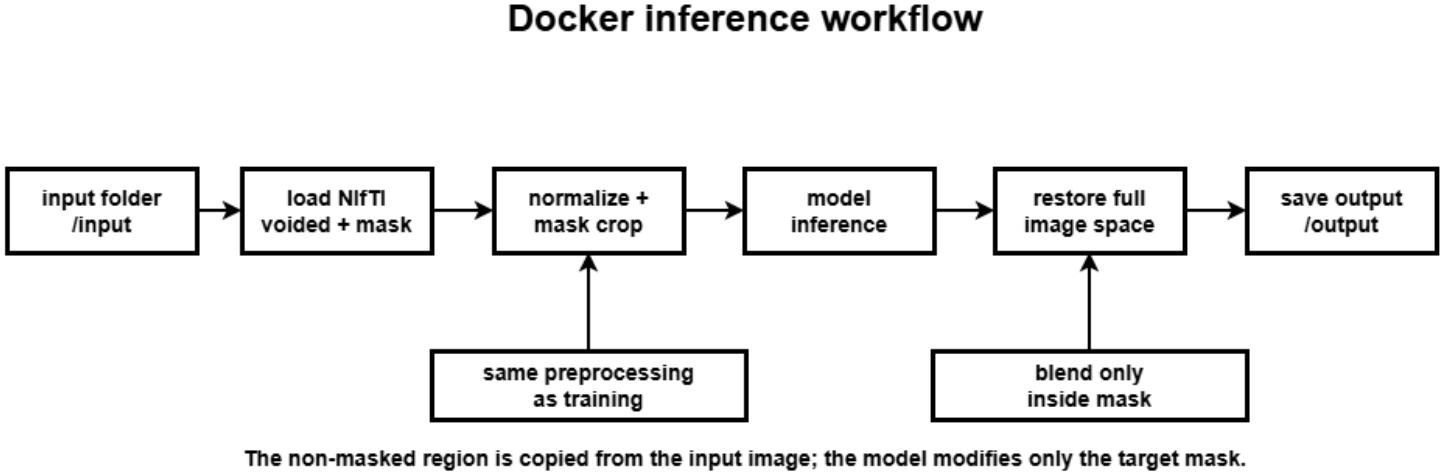
Inference pipeline. The voided image and mask are preprocessed and cropped, passed through the trained generator, and restored to the original image space. Non-masked voxels are retained from the input before the full-resolution NIfTI prediction is written.

## 4 Results

### 4.1 Experimental Setup

Because official validation and test targets are hidden, we report local held-out results using the deterministic split described in Sect. 3.2 (seed 2023, 1251 training and 219 validation cases). Metrics are computed in the inpainting mask region using SSIM, PSNR, MSE, and RMSE. We compare our model with the baseline AutoEncoder and Pix2Pix3D models supplied by the challenge organizers, trained and evaluated under the same split and preprocessing for comparability.

### 4.2 Local Held-Out Performance

Table 1 reports local held-out results. The proposed AnatomyAwareGAN3D improves substantially over the AutoEncoder baseline on all four metrics. Pix2Pix3D remains the strongest local baseline in our experiments; AnatomyAwareGAN3D narrows the gap to Pix2Pix3D relative to the AutoEncoder but does not surpass it under either checkpoint-selection criterion.

### 4.3 Training Observations and Missing Ablations

We report the following as qualitative observations from the development process, not as a controlled ablation study, and we are explicit that they do not by themselves demonstrate the contribution of any individual component: reconstruction-heavy Stage-1 training was more numerically stable than introducing the adversarial term from initialization; the small Stage-2 adversarial weight did not visibly destabilize the reconstruction metrics; and the two checkpoint-selection criteria in Table 1 produce similar but non-identical trade-offs, with the SSIM-selected checkpoint marginally ahead on SSIM and PSNR and the loss-selected checkpoint marginally ahead on MSE and RMSE.

To properly attribute the reported improvement over the AutoEncoder base-line to specific design choices, two controlled ablations are required and were not completed in time for this submission: (1) removing the mirrored-context channel flip(*I*_*v*_) from the generator input while holding all other components and losses fixed, to test whether the symmetry prior in Sect. 2.4 provides measurable benefit; and (2) training with a single-stage schedule (fixed loss weights, no checkpoint-criterion switch) to test whether the two-stage protocol of Sect. 3.8 is responsible for part of the observed gain, as opposed to the architectural changes alone. Both are direct, low-cost re-runs of the existing training pipeline and are the priority next step for this work; without them, the relative contribution of the mirrored-context channel, the attention-modulated skip fusion, and the two-stage schedule to the results in Table 1 remains unverified, and we do not claim otherwise.

### 4.4 Official Challenge Evaluation

The Dockerized model (Sect. 3.9) was submitted to the BraTS 2026 Task 4 (Inpainting) evaluation on the Synapse challenge platform, and the server returned the scores in Table 2, computed over 219 evaluated cases. These are the values returned by the server; we did not recompute them, and the metric definitions follow the organizers’ evaluation code.

**Table 2.** Official server-side scores of the submitted AnatomyAwareGAN3D container (219 evaluated cases). Values are means over cases; *±* gives the per-case standard deviation returned by the server.

| Metric | Value |
|---|---|
| SSIM ↑ | 0.6579 ± 0.1320 |
| PSNR ↑ | 14.8116 ± 4.1975 |
| MSE ↓ | 0.0502 ± 0.0479 |
| RMSE ↓ | 0.0959 |
| MAE ↓ | 0.0546 |
| MSLE ↓ | 0.0236 |

The official scores are clearly lower than the local held-out scores of the same model in Table 1 (SSIM 0.658 versus 0.733–0.734, PSNR 14.81 versus 18.64–18.78, MSE 0.0502 versus 0.0143–0.0147). We also note that the server’s RMSE (0.0959) is not the square root of its mean MSE (approximately 0.224), so the server’s per-metric aggregation differs from a simple transformation of the mean MSE; we report the values as returned. The official scores of the challenge’s AutoEncoder and Pix2Pix3D baselines on the same cases are not available to us, so we make no claim about the model’s rank relative to them on the official data.

### 4.5 Qualitative Prediction Examples

Figures 6 and 7 show full-volume outputs from the submitted AnatomyAware-GAN3D prediction directory. Four cases were selected at evenly spaced positions in the lexicographically sorted list of 219 output filenames, independently of their appearance or evaluation scores. The overview shows orthogonal sections through the center of each volume’s nonzero bounding box; the montage samples nine axial sections within the first selected volume’s nonzero extent. These sections are not selected using the inpainting mask and may intersect preserved tissue as well as synthesized tissue. The available visualization files contain predictions only: without the corresponding voided inputs, masks, or reference targets, the figures cannot localize the synthesized region, establish preservation outside the mask, or demonstrate reconstruction accuracy. They complement the quantitative results by documenting the appearance of the exported volumes.

**Fig. 6.**
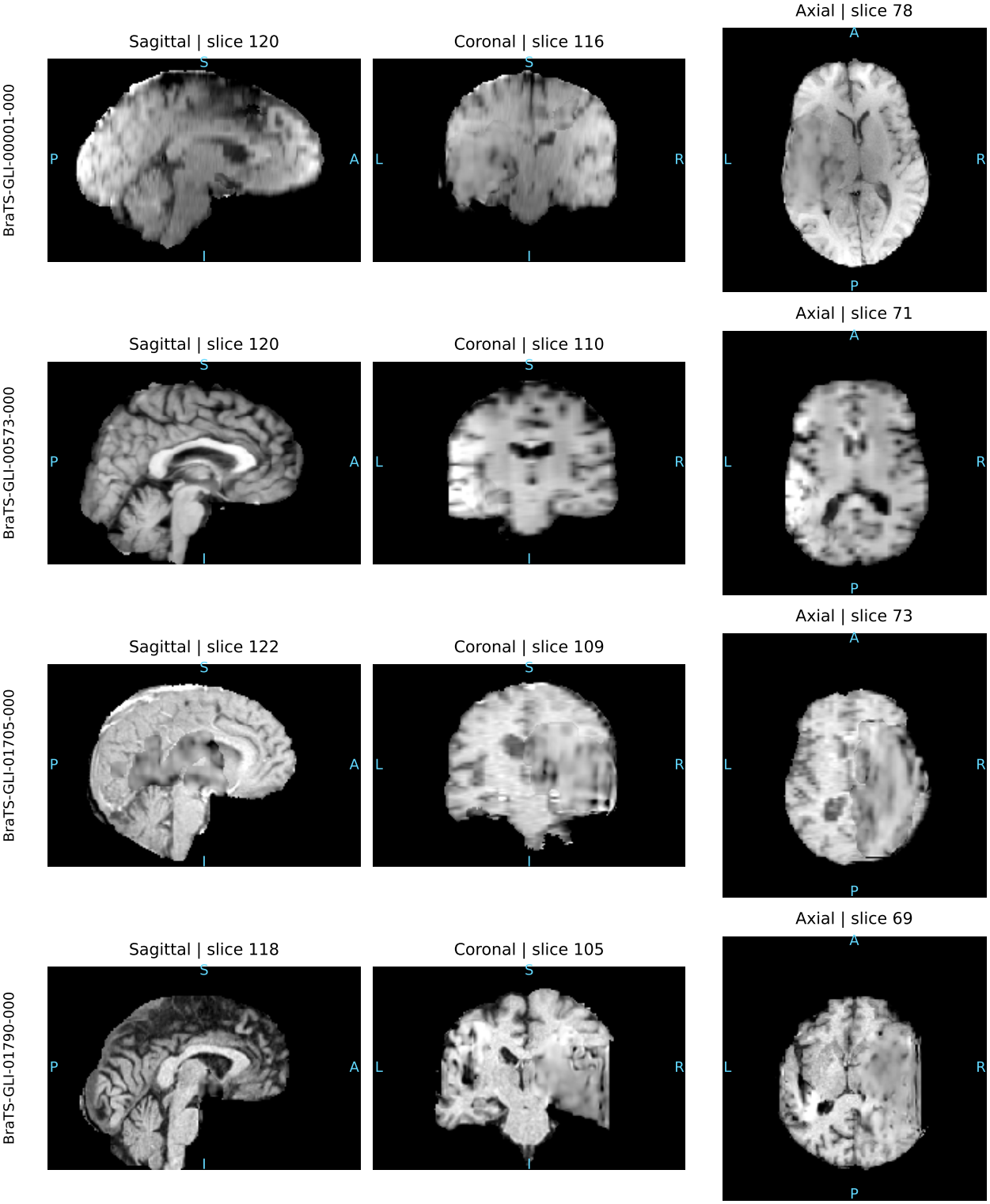
Full-volume prediction examples from AnatomyAwareGAN3D. Rows show cases BraTS-GLI-00001-000, BraTS-GLI-00573-000, BraTS-GLI-01705-000, and BraTS-GLI-01790-000; columns show sagittal, coronal, and axial sections. Volumes are reoriented to RAS+ and displayed in neurological convention. Slice indices are zero-based in the reoriented volume. Display limits use the first and 99th percentiles of finite nonzero intensities separately for each case, shared across its three views. No inpainting-mask overlay or reference target is available.

**Fig. 7.**
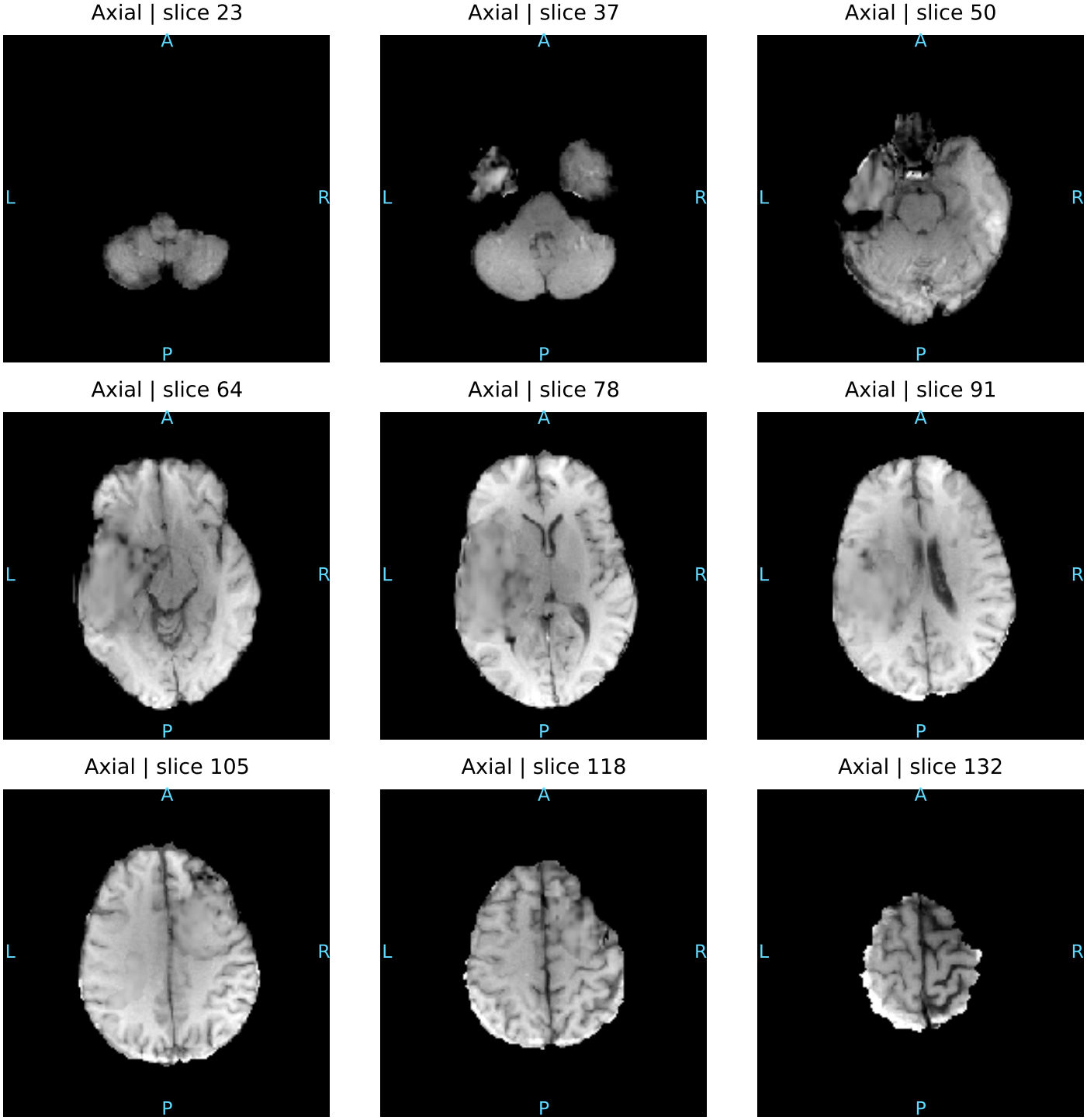
Axial sections of the exported prediction for BraTS-GLI-00001-000. Nine evenly spaced interior sections sample the nonzero axial extent, using the same intensity window throughout. Labels indicate zero-based slice indices after RAS+ reorientation. The montage documents appearance across sections; without masks or reference images, it does not isolate the reconstructed region or establish its fidelity.

## 5 Discussion

The results indicate that the combination of components in AnatomyAware-GAN3D substantially outperforms a simple AutoEncoder baseline on our local split. As noted in Sect. 4.3, however, we cannot currently attribute this improvement to any specific component — the mirrored-context channel, attention-modulated skip fusion, boundary refinement, or the two-stage training schedule — without the ablations described there, and we present the method as a combination whose net effect we measured, not as a set of individually validated contributions.

The method does not exceed the Pix2Pix3D baseline on the local split. Because both methods are evaluated on the identical fixed split described in Sect. 4.1, this gap cannot be attributed to properties of the split itself, since any such effect would apply equally to both models; the more likely explanations are differences in effective model capacity, optimization schedule, or the specific inductive biases each architecture encodes, none of which we have isolated experimentally.

The official server-side result tempers the local findings. On the official cases the model scores a mean SSIM of 0.658 and PSNR of 14.81 dB, well below its local held-out values, which shows that the local split overestimated performance on the official data. We have not isolated the cause. Plausible contributors include differences between the official cases and our local split (for example in mask size, location or scanner characteristics), differences between our local metric implementation and the server’s evaluation code, and overfitting of checkpoint selection to the local split; none of these has been tested here. The official SSIM is numerically closer to our local AutoEncoder result than to the local Pix2Pix3D result, but because the two evaluations differ in cases and possibly in metric implementation, this should not be read as a ranking. Evaluating the baselines on the official cases, or re-scoring our model and the baselines with the server’s metric code on our local split, would resolve this and is a priority for future work.

The reliance on the mirrored-context channel is motivated by documented, approximate bilateral symmetry of the brain [15], but this symmetry is known to be imperfect and can be locally violated by the tumor and surgical or lesion-related tissue changes that motivate the inpainting task in the first place; in cases with substantial mass effect or midline shift, the mirrored channel could plausibly provide misleading rather than helpful context. We flag this as an open question rather than a settled design choice, to be resolved by the ablation in Sect. 4.3 together with a qualitative review of failure cases stratified by mask laterality and size.

A further limitation is the fixed 128 × 128 × 96 crop size (Sect. 3.2), chosen empirically rather than derived from a reported distribution of mask sizes in the training set; very large or irregular masks may exceed the field of view available to the network. Reporting the actual mask-size distribution against the fixed crop, and evaluating performance stratified by mask size, would clarify how often this limitation is encountered in practice and is left for future work alongside the ablations above.

The Dockerized inference pipeline (Sect. 3.9) preserves the non-masked region exactly and modifies only the mask, which reduces the risk of invalid outputs and satisfies the challenge’s one-file-per-case requirement, but is an implementation detail of challenge compliance rather than a scientific contribution, and we do not present it as one.

## 6 Conclusion

We investigated AnatomyAwareGAN3D, a 3D generator for local synthesis of brain MRI tissue in the BraTS 2026 inpainting task, combining gated convolutions, mirrored contralateral context, attention-modulated skip fusion, boundary-aware refinement, multi-scale reconstruction losses, and a local masked-region refiner —each adapted from established prior work rather than proposed as novel individually. On a local held-out split, the combination improves substantially over an AutoEncoder baseline but does not surpass the stronger Pix2Pix3D baseline. On the official challenge server the submitted model obtained a mean SSIM of 0.658 and PSNR of 14.81 dB over 219 cases, below its local held-out performance. We report these outcomes directly rather than overstating it, and we identify the two controlled ablations — removing the mirrored-context channel and removing the two-stage training schedule — required to attribute the observed gain to specific components, together with mask-size-stratified evaluation and multi-fold variability reporting, as concrete next steps before any claim of component-level contribution can be supported.

## Data Availability

The BraTS 2026 dataset used in this study is available through the official BraTS challenge platform, subject to its access and data-use requirements. The processed data, model implementation, and supplementary materials are available from the corresponding author upon reasonable request, subject to applicable dataset restrictions.

https://challenges.synapse.org/Challenges/DetailsPage/Task4?id=syn74274097#Data

## Generative AI Use Statement

Generative AI assistance was used for several writing and documentation tasks, including grammar correction, language refinement, organization of the manuscript, and revision of the manuscript in response to peer review feedback. The scientific method, implementation, experiments, model training, validation outputs, Docker packaging, and final responsibility for the manuscript remain with the authors.

